# Study on the compatibility of lidocaine/prilocaine aerosol with polymer condoms

**DOI:** 10.64898/2026.06.03.26354847

**Authors:** Xiaolu Jiang, Jun Fu, Cuicui Qu, Junhui Huang, Xiaoliang Hu

**Affiliations:** Zhejiang Institute of Medical Device Supervision and Testing, Hangzhou, Zhejiang, China; Shanghai Yunsanova Biotechnology Co.,Ltd, Shanghai, China; NMPA Key Laboratory of Biomedical Optic, Hangzhou, Zhejiang, China

## Abstract

To explore the safety of combined use of lidocaine/prilocaine aerosol and condoms of different materials, this study conducted compatibility tests between them. By observing changes in various physical properties of condom materials after exposure to the aerosol, the compatibility of different polymer materials with the aerosol was analyzed.The results showed that within 15 minutes of exposure to the aerosol, there was no significant difference in all physical properties of natural rubber latex condoms compared with the blank control group (P>0.05), indicating they can be used together. In contrast, obvious changes in physical properties of polyurethane condoms occurred within 5 minutes of exposure (P<0.05), and their performances failed to meet industrial application standards, so combined use is strictly prohibited.This study clarifies the compatibility differences between two mainstream condom materials and lidocaine/prilocaine aerosol, providing experimental evidence and theoretical references for rational matching in clinical and daily use as well as avoiding potential safety risks.

## 1 Introduction

Premature ejaculation is a prevalent sexual dysfunction. It is characterized by persistent or nearly immediate ejaculation immediately after vaginal penetration (lifelong premature ejaculation), or a marked and distressing reduction in ejaculatory latency, typically to less than 3 minutes (acquired premature ejaculation) [1]. It affects males across all age groups. Studies have shown that its prevalence among sexually active men is age-related, with an overall prevalence rate as high as 31% in males aged 18 to 59 years[2]-[5].

Lidocaine/prilocaine aerosol(aerosol) has become a first-line clinical intervention due to its rapid onset and convenient application. Such aerosol reduce glans sensitivity and prolong ejaculatory latency via local anesthetic effects. Their auxiliary ingredients include ethanol, norflurane propellant and polar cosolvents, etc.[6],[7]

Among the common contraceptive methods currently available, condoms offer the highest contraceptive success rate and the lowest incidence of adverse reactions. In addition to preventing conception, correct use of condoms can significantly reduce the risk of HIV and other sexually transmitted infections. As a result, condoms are widely used in clinical practice[8]-[12].

In clinical application, the aerosol inevitably comes into direct contact with condom materials. As a key barrier for contraception and prevention of sexually transmitted diseases, the structural integrity and mechanical properties of condoms directly determine their protective efficacy. There are various types of condom materials with distinctly different chemical properties, while relevant data and literature on their interactions with lidocaine/prilocaine aerosol remain scarce. Most existing studies mainly focus on the anesthetic efficacy, dosage optimization and safety evaluation of local anesthetic preparations[13], or only explore the damaging effects of oily and silicone-based lubricants on condom materials, yet neglect the interactions between aerosols and condoms.

Deterioration in condom properties caused by incompatibility between the aerosol and condom materials will not only reduce contraceptive success rates, but also raise the risk of sexually transmitted disease infection. As a core indicator for evaluating property attenuation after contact between two substances, material compatibility research provides crucial evidence for safe clinical medication and contraceptive protection.

On this basis, this study takes lidocaine/prilocaine aerosol as the research subject to systematically investigate its compatibility with condoms made of common polymer materials. By analyzing the changes in physical properties of materials after contact, the adaptability of condoms of different materials to local anesthetic aerosols is clarified, which can provide experimental support and theoretical references for the rational selection of contraceptive and protective methods in clinical practice and the reduction of potential safety risks for users.

## 2 Materials and Methods

### 2.1 Sample Selection and Information

The aerosol used in this study is Lidocaine and Prilocaine Aerosol.Manufacturers: Shanghai Yunsanova Biotechnology Co.,Ltd.Specification: 5 mL (Each ml of solution contains 150 mg lidocaine and 50 mg prilocaine. ).Batch No.: 251002.

According to public data released by the National Medical Products Administration of China, as of January 15, 2026, there were a total of 87 domestic registration certificates for male condoms in China, including 63 for natural rubber latex(NRL) male condoms, 20 for polyurethane condoms, and 4 for condoms made of other materials. There were 32 imported registration certificates, among which 26 were for NRL male condoms, 4 for polyurethane condoms, and 2 for other material condoms.

Statistical analysis shows that registration certificates of domestic condoms made of NRL and polyurethane account for 95% of the total, while those of imported ones reach 92%. The number of registration certificates reflects the market distribution of condom materials. Given their high market share and representativeness, condoms made of NRL and polyurethane were selected as the research subjects in this study. Detailed information of the test condoms is shown in Table 1.

**Table 1.** Condom Information.

|  | NRL male condoms | Polyurethane condoms |
| --- | --- | --- |
| Brand | Durex | Elephant |
| Model | Nominal width: 52 mm | Smooth surfac, Nominal width: 55 mm ± 2mm |
| Batch No. | 1002873640 | 2412DX19 |

### 2.2 Methods

Due to varying quality levels of commercially available condoms, partial physical property tests were conducted in accordance with ISO 4074-2015 Natural rubber latex male condoms. Requirements and test methods and YY/T 1850-2023 Male condoms. Requirements and test methods for condoms made from polyurethane to verify compliance with standard requirements[14][15].

#### 2.2.1 NRL Male Condoms

NRL Male condoms were divided into three groups, designated as Group 1, Group 1-1 (5-minute aerosol contact) and Group 1-2 (15-minute aerosol contact).

Group 1 was tested in accordance with ISO 4074, and the test scheme is presented in Table 2.

**Table 2.** Sampling inspection scheme of NRL male condoms.

| No. | Items | Clause | Judgment Criteria |
| --- | --- | --- | --- |
| 1 | Bursting volume and pressure | 10 | 315[10, 11] |
| 2 | Holes | 12 | 500[3, 4] |

To reduce sample consumption of Test Group 1-1 and 1-2, ISO 2859-1-2026 *Sampling procedures for inspection by attributes — Part 1: Sampling schemes indexed by acceptance quality limit (AQL) for lot-by-lot inspection* was consulted[16]. The sample code was reduced to K with a sample size of 125 specimens. When the AQL was set at 1.5 for bursting volume and pressure tests, the acceptance number and rejection number were determined as 5 and 6 respectively. When the AQL was 0.25 for pinhole inspection, the acceptance number was 1 and the rejection number was 2.

A total of 270 condoms (20 spares) of Test Group 1-1 were deprived of outer packages. 125 condoms were tested for holes, and another 125 condoms were tested for bursting volume and pressure. Aerosol was sprayed onto each condom to simulate actual use. The inner side was sprayed three times, with about 50 microlitres discharged per actuation. The aerosol contact time was set as 5 minutes, and all samples were tested separately.

A total of 270 condoms (20 spares) of Test Group 1-2 were deprived of outer packages. 125 condoms were tested for holes, and another 125 condoms were tested for bursting volume and pressure. Aerosol was sprayed onto each condom to simulate actual use. The inner side was sprayed three times, with about 50 microlitres discharged per actuation. The aerosol contact time was set as 15 minutes, and all samples were tested separately.

#### 2.2.2 Polyurethane Condoms

Polyurethane condoms were divided into three groups, designated as Control Group 2, Test Group 2-1 (5-minute aerosol contact) and Test Group 2-2 (15-minute aerosol contact).

Group 2 was tested in accordance with YY/T 1850, and the test scheme is presented in Table 3.]

**Table 3.** Sampling inspection scheme of polyurethane condoms.

| No. | Items | Clause | Judgment Criteria |
| --- | --- | --- | --- |
| 1 | Bursting volume and pressure | 9.4 | 315[10, 11] |
| 2 | Force and elongation at break | 9.5 | 13[0, 1] |
| 3 | Holes | 11 | 500[3, 4] |

To reduce sample consumption of Test Group 2-1 and 2-2, ISO 2859-1 was consulted. The sample code was reduced to K with a sample size of 125 specimens. When the AQL was set at 1.5 for bursting volume and pressure tests, the acceptance number and rejection number were determined as 5 and 6 respectively. When the AQL was 0.25 for pinhole inspection, the acceptance number was 1 and the rejection number was 2.

A total of 283 condoms (20 spares) of Test Group 2-1 were deprived of outer packages. 125 condoms were tested for holes, 125 condoms were tested for bursting volume and pressure, and 13 samples were tested for force and elongation at break . Aerosol was sprayed onto each condom to simulate actual use. The inner side was sprayed three times, with about 50 microlitres discharged per actuation. The aerosol contact time was set as 5 minutes, and all samples were tested separately.

A total of 283 condoms (20 spares) of Test Group 2-2 were deprived of outer packages. 125 condoms were tested for holes, 125 condoms were tested for bursting volume and pressure, and 13 samples were tested for force and elongation at break . Aerosol was sprayed onto each condom to simulate actual use. The inner side was sprayed three times, with about 50 microlitres discharged per actuation. The aerosol contact time was set as 15 minutes, and all samples were tested separately.

## 3 Results

### 3.1 NRL Male Condoms

Test results of bursting volume and pressure for NRL male condoms in Group 1 are shown in Table 4.

**Table 4.**
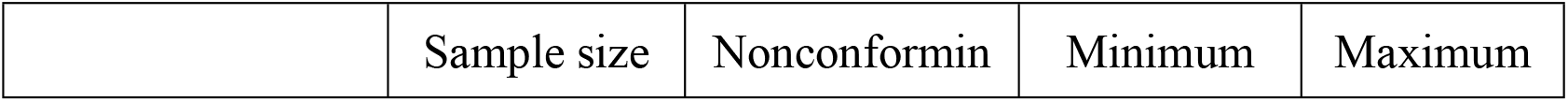
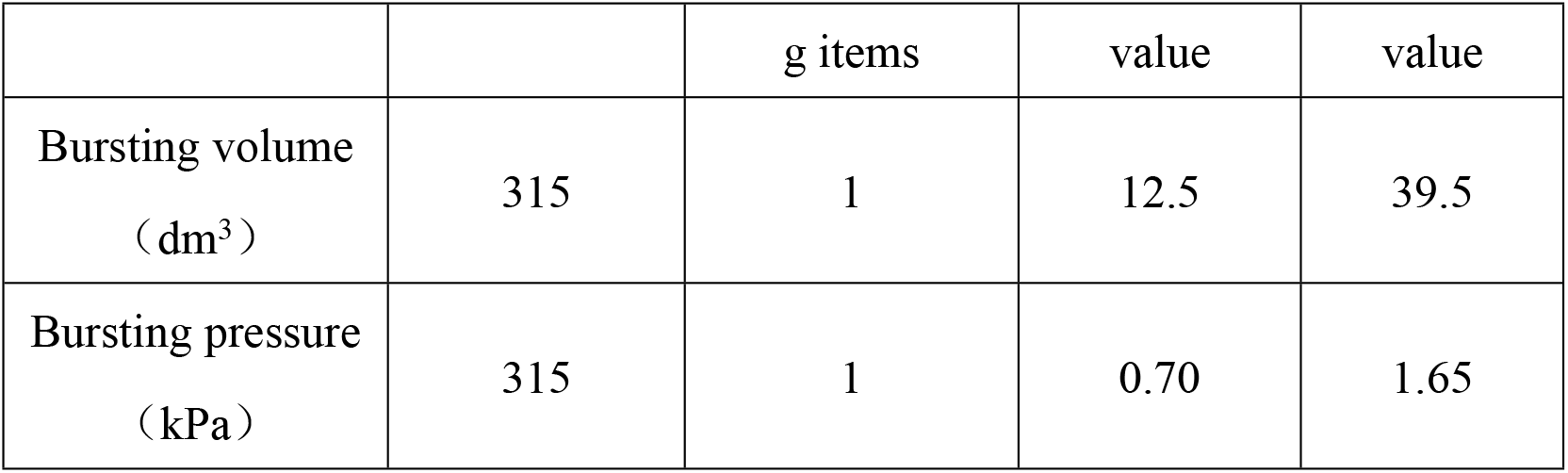
Test results of bursting volume and pressure for the Group 1.

Test results of holes for NRL male condoms in Group 1 are shown in Table 5.

**Table 5.**
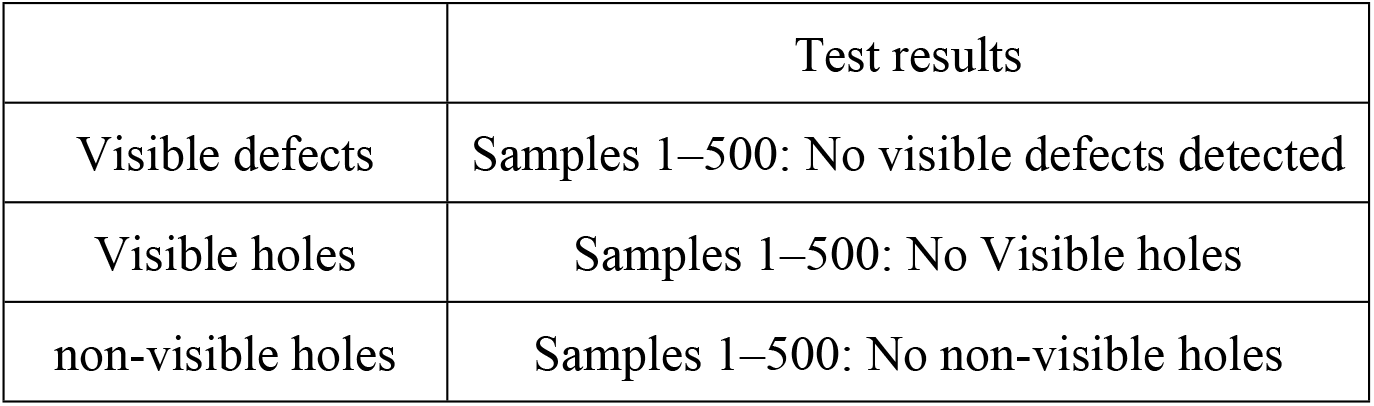
Test results of holes for the Group 1.

|  | Test results |
| --- | --- |
| Visible defects | Samples 1–500: No visible defects detected |
| Visible holes | Samples 1–500: No Visible holes |
| non-visible holes | Samples 1–500: No non-visible holes |

Test results of bursting volume and pressure for NRL male condoms in Group 1-1 are shown in Table 6.

**Table 6.** Test results of bursting volume and pressure for the Group 1-1.

|  | Sample size | Nonconforming items | Minimum value | Maximum value |
| --- | --- | --- | --- | --- |
| Bursting volume (dm<sup>3</sup>) | 125 | 0 | 23.5 | 40.0 |
| Bursting pressure (kPa) | 125 | 0 | 1.00 | 1.65 |

Test results of holes for NRL male condoms in Group 1-1 are shown in Table 7.

**Table 7.** Test results of holes for the Group 1-1.

|  | Test results |
| --- | --- |
| Visible defects | Samples 1–125: No visible defects detected |
| Visible holes | Samples 1–125: No Visible holes |
| non-visible holes | Samples 1–125: No non-visible holes |

Test results of bursting volume and pressure for NRL male condoms in Group 1-2 are shown in Table 8.

**Table 8.** Test results of bursting volume and pressure for the Group 1.

|  | Sample size | Nonconforming items | Minimum value | Maximum value |
| --- | --- | --- | --- | --- |
| Bursting volume (dm<sup>3</sup>) | 125 | 0 | 22.0 | 41.5 |
| Bursting pressure (kPa) | 125 | 0 | 1.05 | 1.65 |

Test results of holes for NRL male condoms in Group 1-2 are shown in Table 9.

**Table 9.**
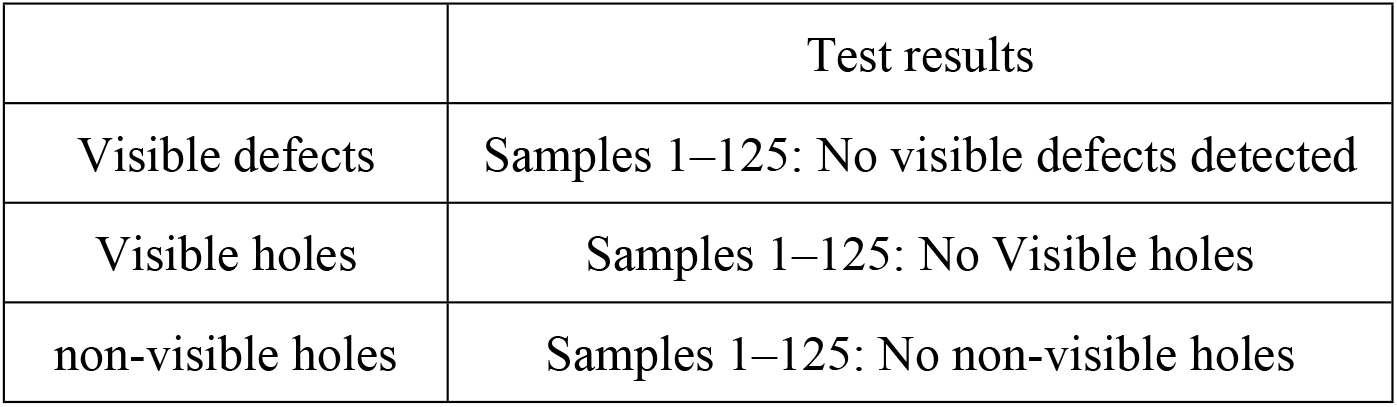
Test results of holes for the Group 1-2.

### 3.2 Polyurethane Condoms

Test results of bursting volume and pressure for polyurethane condoms in Group 2 are shown in Table 10.

**Table 10.** Test results of bursting volume and pressure for the Group 2.

|  | Sample size | Nonconforming items | Minimum value | Maximum value |
| --- | --- | --- | --- | --- |
| Bursting volume (dm<sup>3</sup>) | 315 | 9 | 0.5 | 12.0 |
| Bursting pressure (kPa) | 315 | 6 | 1.65 | 7.70 |

Test results of holes for polyurethane condoms in Group 2 are shown in Table 11.

**Table 11.** Test results of holes for the Group 2.

|  | Test results |
| --- | --- |
| Visible defects | Samples 1–500: No visible defects detected |
| Visible holes | Samples 1–500: No Visible holes |
| non-visible holes | Samples 37, 72: Leakage; Others: No leakage |

Test results of force and elongation at break for polyurethane condoms in Group 2 are shown in Table 12.

**Table 12.** Test results of force and elongation at break for the Group 2.

| Serial number | Force at break<br>(N) | Elongation at<br>break |
| --- | --- | --- |
| 1 | 22.024 | 570% |
| 2 | 20.500 | 566% |
| 3 | 22.210 | 571% |
| 4 | 28.471 | 600% |
| 5 | 22.423 | 580% |
| 6 | 26.351 | 586% |
| 7 | 20.440 | 561% |
| 8 | 36.513 | 599% |
| 9 | 25.466 | 568% |
| 10 | 22.300 | 548% |
| 11 | 25.215 | 559% |
| 12 | 26.644 | 564% |
| 13 | 22.035 | 541% |

Test results of bursting volume and pressure for polyurethane condoms in Group 2-1 are shown in Table 13.

**Table 13.** Test results of bursting volume and pressure for the Group 2-1.

|  | Sample<br>size | Nonconforming<br>items | Minimum<br>value | Maximum<br>value |
| --- | --- | --- | --- | --- |
| Bursting volume (dm<sup>3</sup>) | 125 | 6 | 2.5 | 10.5 |
| Bursting pressure (kPa) | 125 | 7 | 1.55 | 5.30 |

Test results of holes for polyurethane condoms in Group 2-1 are shown in Table 14.

**Table 14.** Test results of holes for the Group 2-1.

|  | Test results |
| --- | --- |
| Visible defects | Samples 1–125: No visible defects detected |
| Visible holes | Samples 1–125: No Visible holes |
| non-visible holes | Samples 26,34,61,69,88,95,103: Leakage;<br>Others: No leakage |

Test results of force and elongation at break for polyurethane condoms in Group 2-1 are shown in Table **Error! Reference source not found**.15.

**Table 15.** Test results of force and elongation at break for the Group 2-1.

| Serial number | Force at break<br>(N) | Elongation at<br>break |
| --- | --- | --- |
| 1 | 9.777 | 465% |
| 2 | 6.692 | 405% |
| 3 | 7.957 | 409% |
| 4 | 7.009 | 362% |
| 5 | 8.764 | 435% |
| 6 | 3.878 | 231% |
| 7 | 10.074 | 455% |
| 8 | 7.614 | 418% |
| 9 | 8.207 | 442% |
| 10 | 6.777 | 383% |
| 11 | 8.104 | 419% |
| 12 | 5.099 | 310% |
| 13 | 7.107 | 409% |

Test results of bursting volume and pressure for polyurethane condoms in Group 2-2 are shown in Table 16.

**Table 16.** Test results of bursting volume and pressure for the Group 2-2.

|  | Sample size | Nonconforming items | Minimum value | Maximum value |
| --- | --- | --- | --- | --- |
| Bursting volume (dm<sup>3</sup>) | 125 | 9 | 2.0 | 10.5 |
| Bursting pressure (kPa) | 125 | 12 | 1.70 | 4.70 |

Test results of holes for polyurethane condoms in Group 2-2 are shown in Table 17.

**Table 17.** Test results of holes for the group 2-2.

|  | Test results |
| --- | --- |
| Visible defects | Samples 1–125: No visible defects detected |
| Visible holes | Samples 1–125: No Visible holes |
| non-visible holes | Samples 14,37,59,84,92: Leakage; Others: No leakage |

Test results of force and elongation at break for polyurethane condoms in Group 2-2 are shown in Table 18.

**Table 18.**
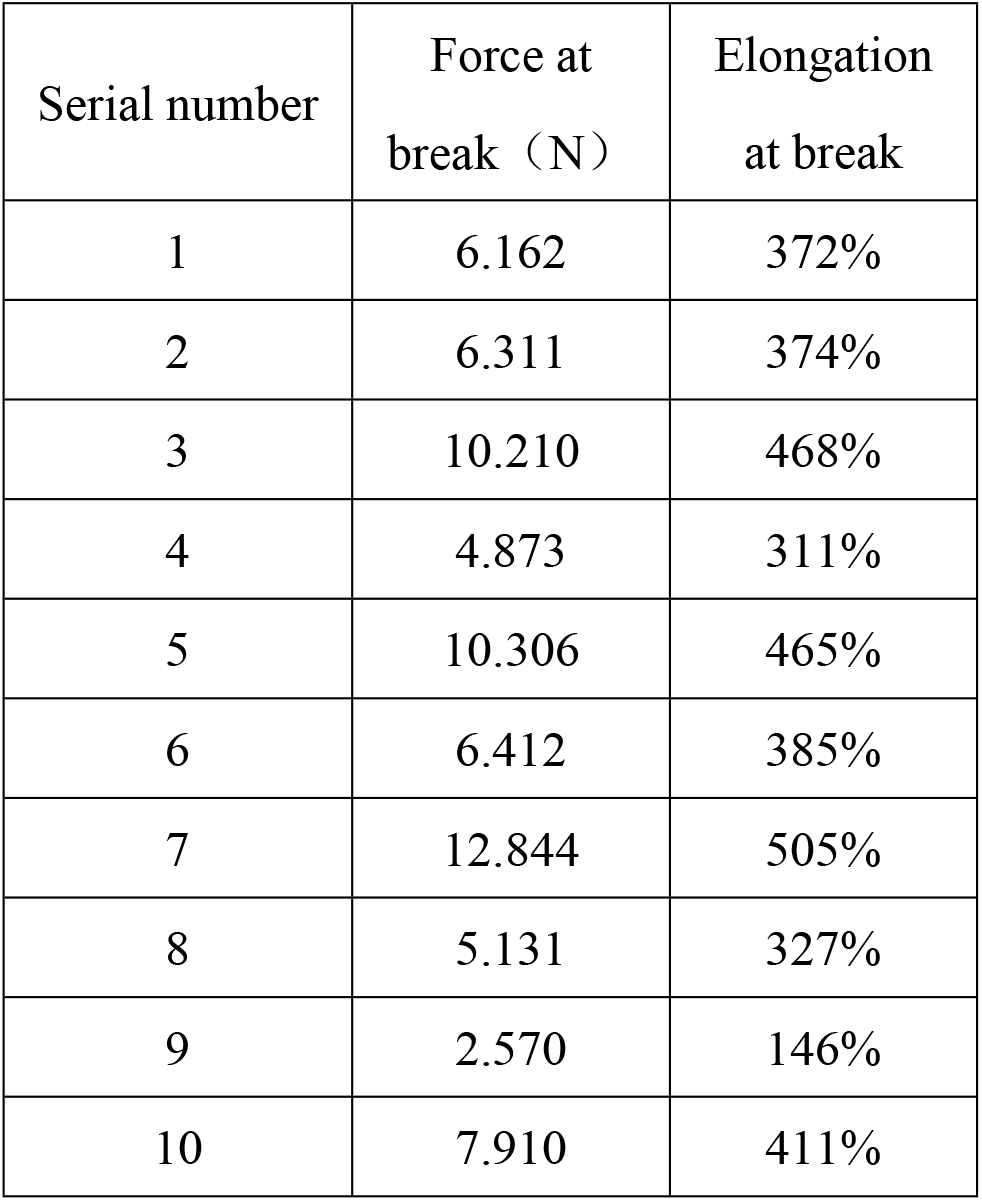
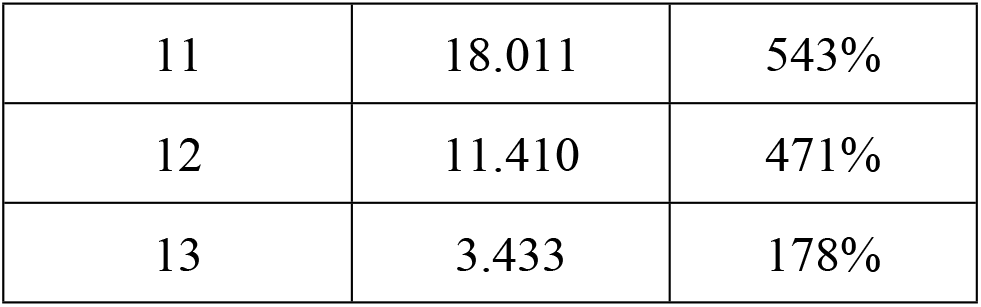
Test results of force and elongation at break for the Group 2-2.

| Serial number | Force at break (N) | Elongation at break |
| --- | --- | --- |
| 1 | 6.162 | 372% |
| 2 | 6.311 | 374% |
| 3 | 10.210 | 468% |
| 4 | 4.873 | 311% |
| 5 | 10.306 | 465% |
| 6 | 6.412 | 385% |
| 7 | 12.844 | 505% |
| 8 | 5.131 | 327% |
| 9 | 2.570 | 146% |
| 10 | 7.910 | 411% |
| 11 | 18.011 | 543% |
| 12 | 11.410 | 471% |
| 13 | 3.433 | 178% |

## 4 Conclusion

### 4.1 NRL Male Condoms

The test results of bursting volume, bursting pressure and holes for NRL male condoms in Group 1 show that all test specimens comply with ISO 4074.

S_d_ was calculated using Equation (1) specified in GB/T 4889-2008 *Statistical interpretation of data. Techniques of estimation and tests relating to means and variances of normal distribution*. S_d_ refers to the combined standard deviation of the two groups of data [17].

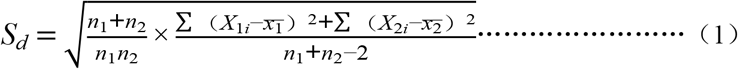

If 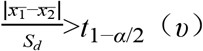, reject the hypothesis that the two means are equal.

Where

*n*_*1*_ is sample size of the first group;*n*_*2*_ is sample size of the second group; *X*_*1i*_ is each individual value in the first group; 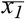 is Average of the first group; *X*_*2i*_ is each individual value in the second group;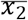 is Average of the second group; *α* is significance level, set to 0.05; *t*_1-α/2_*(υ)*is the 1-*α*/2 quantile of the t-distribution with degrees of freedom.

#### 4.1.1 Bursting Volume Data Analysis

Statistical analysis was performed on the test sample data, as shown in Table 19.

**Table 19.**
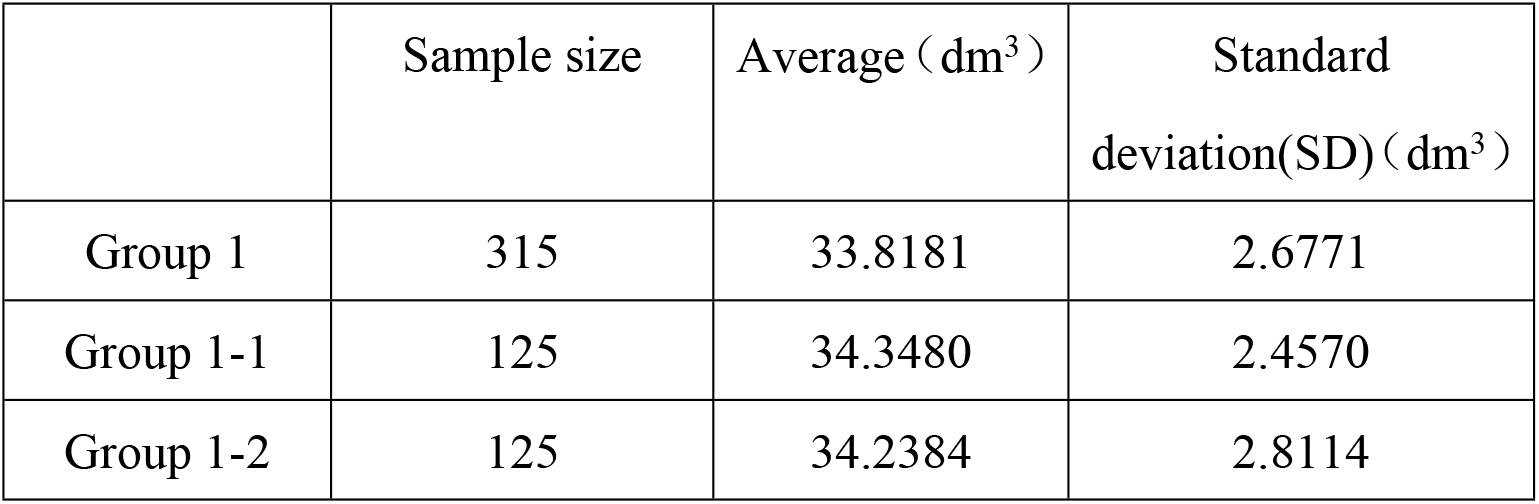
Analysis of bursting volume data.

All samples from Groups 1-1 and 1-2 passed the bursting volume test and met the requirements of ISO 4074.

The *S*_*d*_ of Groups 1 and 1-1 was calculated to be 0.2766.According to the table in Appendix A of GB/T 4889-2008, *t*_*0*.*975*_*(500)*=1.9647.

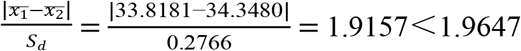, No statistically significant difference was found between the means of the two groups (*P* > 0.05).

The *S*_*d*_ of Groups 1 and 1-2 was calculated to be 0.2871.According to the table in Appendix A of GB/T 4889-2008, *t*_*0*.*975*_*(500)*=1.9647.

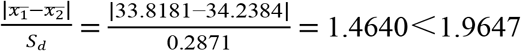, No statistically significant difference was found between the means of the two groups (*P* > 0.05).

#### 4.1.2 Bursting Pressure Data Analysis

Statistical analysis was performed on the test sample data, as shown in Table 20.

**Table 20.**
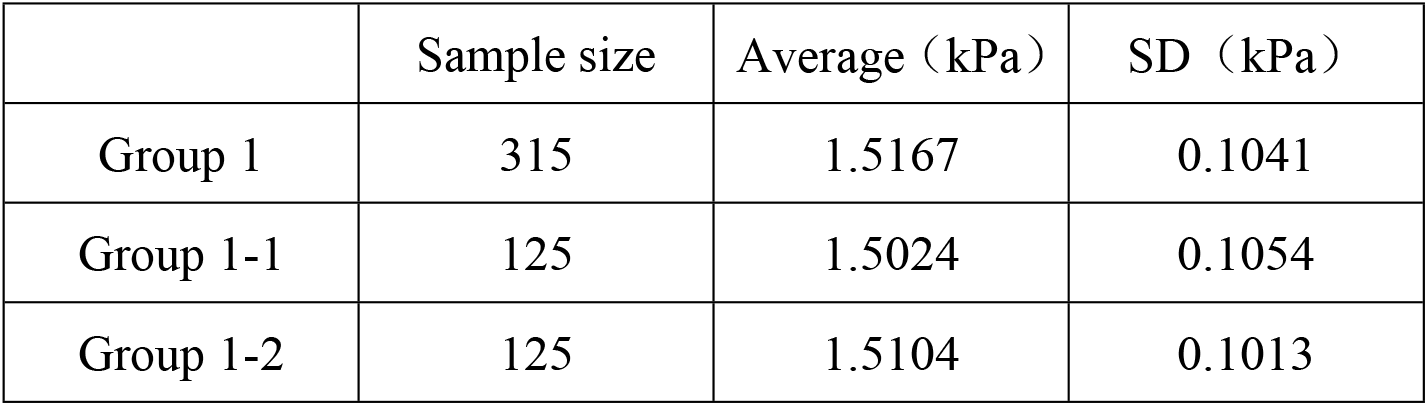
Analysis of bursting pressure data.

All samples from Groups 1-1 and 1-2 passed the bursting pressure test and met the requirements of ISO 4074.

The *S*_*d*_ of Groups 1 and 1-1 was calculated to be 0.0110.According to the table in Appendix A of GB/T 4889-2008, *t*_*0*.*975*_*(500)*=1.9647.

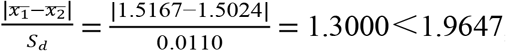, No statistically significant difference was found between the means of the two groups (*P* > 0.05).

The *S*_*d*_ of Groups 1 and 1-2 was calculated to be 0.0109.According to the table in Appendix A of GB/T 4889-2008, *t*_*0*.*975*_*(500)*=1.9647.

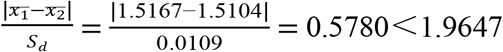, No statistically significant difference was found between the means of the two groups (*P* > 0.05).

#### 4.1.3 Holes Data Analysis

No nonconforming items were detected in the holes test for Groups 1-1 and 1-2 and met the requirements of ISO 4074.

All specimens passed the holes test for Groups 1, 1-1 and 1-2, and consistent results were obtained between the two groups.

### 4.2 Polyurethane Condoms

The test results of bursting volume, bursting pressure, holes, and force and elongation at break for polyurethane condoms in Group 2 show that all test specimens comply with YY/T 1850.

#### 4.2.1 Bursting Volume Data Analysis

Statistical analysis was performed on the test sample data, as shown in Table 21.

**Table 21.** Analysis of bursting volume data.

|  | Sample size | Average (dm<sup>3</sup>) | SD (dm<sup>3</sup>) |
| --- | --- | --- | --- |
| Group 2 | 315 | 8.4794 | 1.8091 |
| Group 2-1 | 125 | 7.6452 | 1.6689 |
| Group 2-2 | 125 | 7.5080 | 1.6172 |

The *S*_*d*_ of Groups 2 and 2-1 was calculated to be 0.1872.According to the table in Appendix A of GB/T 4889-2008, *t*_*0*.*975*_*(500)*=1.9647.

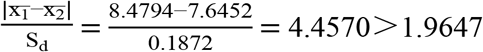, A statistically significant difference was found between the two group means (P < 0.05), and the null hypothesis of equal means was rejected. 6 nonconforming items were detected in the bursting volume test of Group 2-1, which failed to meet the requirements of YY/T 1850-2023.

The *S*_*d*_ of Groups 2 and 2-2 was calculated to be 0.1857.According to the table in Appendix A of GB/T 4889-2008, *t*_*0*.*975*_*(500)*=1.9647.

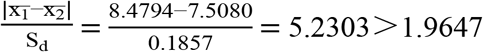, A statistically significant difference was found between the two group means (P < 0.05), and the null hypothesis of equal means was rejected. 9 nonconforming items were detected in the bursting volume test of Group 2-1, which failed to meet the requirements of YY/T 1850.

### 4.2.2 Bursting Pressure Data Analysis

Statistical analysis was performed on the test sample data, as shown in Table 22.

**Table 22.** Analysis of bursting pressure data.

|  | Sample size | Average (kPa) | SD (kPa) |
| --- | --- | --- | --- |
| Group 2 | 315 | 3.3683 | 0.9061 |
| Group 2-1 | 125 | 2.9432 | 0.6653 |
| Group 2-2 | 125 | 2.9344 | 0.6641 |

The *S*_*d*_ of Groups 2 and 2-1 was calculated to be 0.0893.According to the table in Appendix A of GB/T 4889-2008, *t*_*0*.*975*_*(500)*=1.9647.

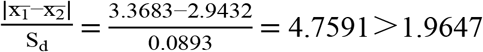, A statistically significant difference was found between the two group means (P < 0.05), and the null hypothesis of equal means was rejected. 7 nonconforming items were detected in the bursting volume test of Group 2-1, which failed to meet the requirements of YY/T 1850-2023.

The *S*_*d*_ of Groups 2 and 2-1 was calculated to be 0.0893.According to the table in Appendix A of GB/T 4889-2008, *t*_*0*.*975*_*(500)*=1.9647.

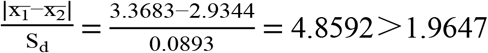, A statistically significant difference was found between the two group means (P < 0.05), and the null hypothesis of equal means was rejected. 12 nonconforming items were detected in the bursting volume test of Group 2-2, which failed to meet the requirements of YY/T 1850-2023.

#### 4.2.3 Holes Data Analysis

Statistical analysis was performed on the test sample data, as shown in Table 23.

**Table 23.**
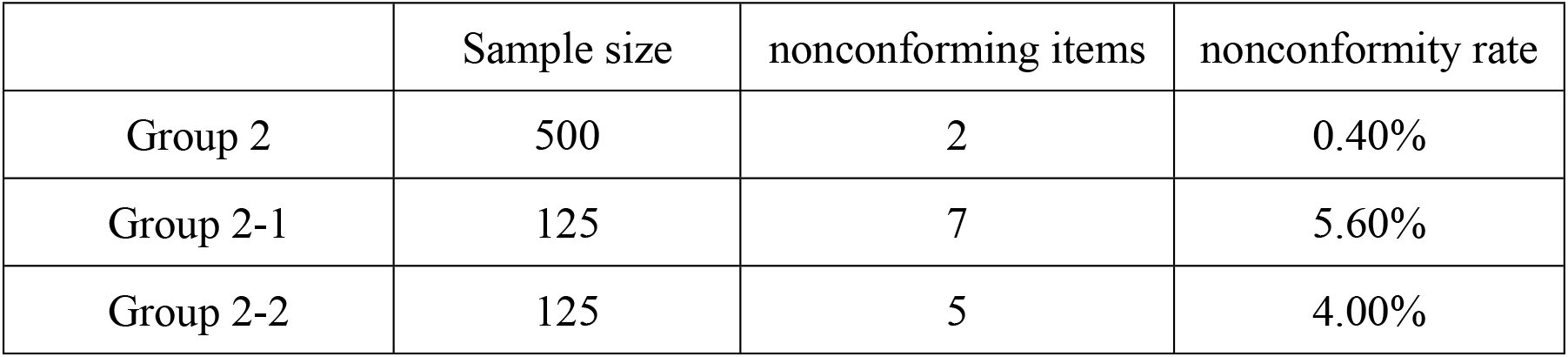
Analysis of holes data.

Analysis of nonconformity rates for Group 2 and Group 2-1 revealed 7 nonconforming items in the holes test of Group 2-1,which failed to meet the requirements. The nonconformity rate of Test Group 2-1 was 14 times that of Group 2, and a significant difference was observed between the two groups.

Analysis of nonconformity rates for Group 2 and Group 2-2 revealed 5 nonconforming items in the holes test of Group 2-2,which failed to meet the requirements. The nonconformity rate of Test Group 2-2 was 10 times that of Group 2, and a significant difference was observed between the two groups.

#### 4.2.4 Analysis of Force and Elongation at Break Data

Statistical analysis was performed on the test sample data, as shown in Table 24.

**Table 24.** Analysis of force at break data.

|  | Sample size | Average (N) | SD (N) |
| --- | --- | --- | --- |
| Group 2 | 13 | 24.6609 | 4.3618 |
| Group 2-1 | 13 | 7.4661 | 1.7062 |
| Group 2-2 | 13 | 8.1218 | 4.3054 |

The *S*_*d*_ of Groups 2 and 2-1 was calculated to be 1.2990.According to the table in Appendix A of GB/T 4889-2008, *t*_*0*.*975*_*(24)*=2.0639.

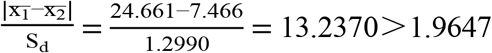, A statistically significant difference was found between the two group means (P < 0.05), and the null hypothesis of equal means was rejected. 13 nonconforming items were detected in the force at break test of Group 2-1, which failed to meet the requirements of YY/T 1850-2023.

The *S*_*d*_ of Groups 2 and 2-2 was calculated to be 1.6998.According to the table in Appendix A of GB/T 4889-2008, *t*_*0*.*975*_*(24)*=2.0639.

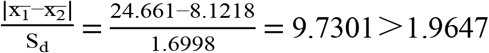, A statistically significant difference was found between the two group means (P < 0.05), and the null hypothesis of equal means was rejected. 13 nonconforming items were detected in the force at break test of Group 2-2, which failed to meet the requirements of YY/T 1850-2023.

#### 4.2.5 Analysis of Elongation at Break Data

Statistical analysis was performed on the test sample data, as shown in Table 25.

**Table 25.** Analysis of elongation at break data.

|  | Sample size | Average | SD |
| --- | --- | --- | --- |
| Group 2 | 13 | 5.7023 | 0.1751 |
| Group 2-1 | 13 | 3.9562 | 0.6407 |
| Group 2-2 | 13 | 3.8123 | 1.1890 |

The *S*_*d*_ of Groups 2 and 2-1 was calculated to be 0.1842.According to the table in Appendix A of GB/T 4889-2008, *t*_*0*.*975*_*(24)*=2.0639.

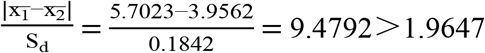, A statistically significant difference was found between the two group means (P < 0.05), and the null hypothesis of equal means was rejected. 13 nonconforming items were detected in the force at break test of Group 2-1, which failed to meet the requirements of YY/T 1850-2023.

The *S*_*d*_ of Groups 2 and 2-2 was calculated to be 0.3333.According to the table in Appendix A of GB/T 4889-2008, *t*_*0*.*975*_*(24)*=2.0639.

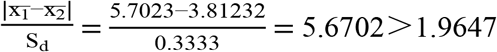, A statistically significant difference was found between the two group means (P < 0.05), and the null hypothesis of equal means was rejected. 13 nonconforming items were detected in the force at break test of Group 2-2, which failed to meet the requirements of YY/T 1850-2023.

## 5 Conclusions

In this study, no statistically significant differences in physical properties were found for NRL male condoms between the group without contact and the group with an aerosol contact time within 15 minutes (P > 0.05). It is indicated that the lidocaine/prilocaine aerosol exerts no significant impact on their physical properties, and the aerosol can be used together with NRL male condoms.

Statistically significant differences in physical properties were observed for polyurethane condoms between the non-contact group and the group with an aerosol contact time within 5 minutes (P < 0.05). This indicates that lidocaine/prilocaine aerosol has a notable adverse effect and degrades the physical performance of polyurethane condoms. Since the deteriorated properties no longer comply with relevant industrial standards, this aerosol shall not be used together with polyurethane condoms.

## Data Availability

The minimal data set [and accompanying code, where generated] is available at [repository name] via [DOI hyperlink] No data was generated by this study. The following existing data sources were used: [dataset name] from [dataset location] available via [web address or DOI hyperlink]. For Study Protocols: No datasets were generated or analysed during the current study. All relevant data from this study will be made available upon study completion.

